# Exploring how an e-cigarette intervention influenced tobacco smoking behaviour in people accessing homelessness services: findings from the SCeTCH trial process evaluation

**DOI:** 10.1101/2025.02.17.25322409

**Authors:** Allison Ford, Lauren McMillan, Sharon Cox, Kirstie Soar, Francesca Pesola, Caitlin Notley, Rachel Brown, Emma Ward, Bethany Gardner, Anna Varley, Charlotte Mair, Jessica Lennon, Janine Brierley, Amy Edwards, Danielle Micthell, Debbie Robson, Peter Hajek, Allan Tyler, Steve Parrott, Jinshuo Li, Linda Bauld, Lynne Dawkins

## Abstract

**Background:** Smoking prevalence among people experiencing homelessness is high. This study explored the factors which influenced potential smoking abstinence among participants receiving an e-cigarette (EC) intervention within the Smoking Cessation Trial in Centres for Homelessness (SCeTCH) across Great Britian.

**Methods:** Using the Capability Opportunity Motivation – Behaviour (COM-B) model, hypothesised and emerging mediators were explored quantitatively via measures in baseline and follow-up questionnaires (n=239) and qualitatively via interviews with homelessness centre staff (n=16) and participants (n=31) who received an EC starter pack and 4-week e-liquid supply. We purposively sampled 8 centres for interviews, representing varied regions and participant vaping/smoking and sociodemographic status. Quantitative measures were analysed descriptively. Qualitative data were analysed thematically. Data from the two approaches were mapped onto the COM-B for combined analysis and reporting.

**Results:** After receiving the intervention, participants demonstrated high capability to use EC and appreciation of EC for harm reduction. Some participants reported dissatisfaction with vaping. Opportunity for behaviour change was strengthened by perceived acceptability to vape and social support beyond the centre but hindered by a smoking culture and perceived lack of staff support. Motivation was enhanced by EC efficacy belief, pride from cutting down, and financial benefits of vaping, but negatively impacted by challenging personal circumstances.

**Conclusion:** For people accessing homelessness support centres, smoking abstinence remains challenging. More intensive support and a variety of approaches to support smoking cessation, particularly those which address the psychosocial factors which hinder smoking abstinence, may be required. Future research should focus on how this can be achieved.

## INTRODUCTION

Tobacco smoking is a key driver of health inequalities with disproportionately high prevalence and smoking-related morbidity and death among disadvantaged populations (Hosseinpoor et al., 2012; Lynch et al., 1997). Among the United Kingdom (UK) general population, 11.9% of adults smoked cigarettes in 2023, compared with 20.2% in 2011 (Office for National Statistics [ONS], 2024). A key driver of this downward trend in smoking prevalence is the implementation of a comprehensive package of population-and individual-level tobacco control measures introduced to support smoking cessation and reduce uptake (Flor et al., 2021). Yet smoking prevalence among people accessing homelessness support services remains substantially high, with estimates around 76% in the UK (Homeless Link, 2022). High levels of smoking, greater tobacco dependence compared with the general population, and engagement with potentially risky behaviours (i.e. smoking unfiltered cigarettes, discarded butts and illicit tobacco), may exacerbate already high levels of poor respiratory health among this group (Aloot et al., 1993; Garner & Ratschen, 2013; Chen et al., 2016).

People experiencing homelessness report desire and motivation to quit (Garner & Ratschen, 2013; Okuyemi et al., 2013; Porter at al., 2017; Dawkins et al., 2019) yet are less likely to access free UK National Health Service (NHS) Stop Smoking Services (SSS) (Dawkins et al., 2019). Barriers to accessing smoking cessation support are complex. There is a misperception among health practitioners that people experiencing homelessness do not want to quit, concomitant with this group reporting lack of support or active discouragement to quit (Garner & Ratschen, 2013). Homeless shelter staff report a lack of knowledge of local cessation resources, feel unqualified to provide advice, and do not consider cessation a priority for residents (Porter et al., 2017). Other barriers include high levels of boredom and stress which are difficult to cope with when craving nicotine (Okuyemi et al., 2006), smoking as a coping mechanism (Stewart et al., 2015), low risk perception of smoking compared to other behaviours (Garner & Ratschen, 2013), smoking combined with substance use (Okuyemi et al., 2006), and scepticism over the effectiveness of traditional methods (Pratt et al., 2019). Social and environmental barriers include the ubiquity of smoking in homelessness settings (Stewart et al., 2015), peer influence (Garner & Ratschen, 2013), and the value of smoking as a means of socialisation (Porter et al., 2017; Pratt et al., 2019).

There is high certainty evidence from randomised controlled trials (RCTs) that nicotine-containing e-cigarettes (EC) increase quit rates compared to nicotine replacement therapy (NRT) (Lindson et al., 2024). While there has previously been inconclusive evidence on the effectiveness of EC for reducing tobacco use in people experiencing homelessness (Vijayaraghavan et al., 2020), EC may be a useful tool given traditional stop smoking methods are not as effective for this group as general populations (Lindson et al., 2024; Vijayaraghavan et al., 2020).

Based on a feasibility study (Cox et al., 2021; Dawkins et al., 2020), the Smoking Cessation Trial in Centres for Homelessness (SCeTCH) compared the effects of EC versus usual care for smoking cessation when provided at homelessness support centres across Great Britain (England, Scotland, Wales) (Figure 1) (Cox et al., 2022). Providing an EC starter kit was not an effective intervention for sustained abstinence at 24-weeks, however, significant reductions in tobacco smoking and differences between arms in short-term quitting were reported (Figure 1) (Dawkins et al., 2024). This paper reports results from the embedded process evaluation within SCeTCH, which aimed to explore the mechanisms through which the EC intervention influenced changes in smoking behaviour. The processes that lead to changes in smoking behaviour among people experiencing or at-risk of homelessness are not widely understood. This paper aims to fill this gap by exploring hypothesised and emerging (i.e. inductively derived) mediators which served as barriers and facilitators to smoking reduction and short-term abstinence during the trial.

**Figure 1:**
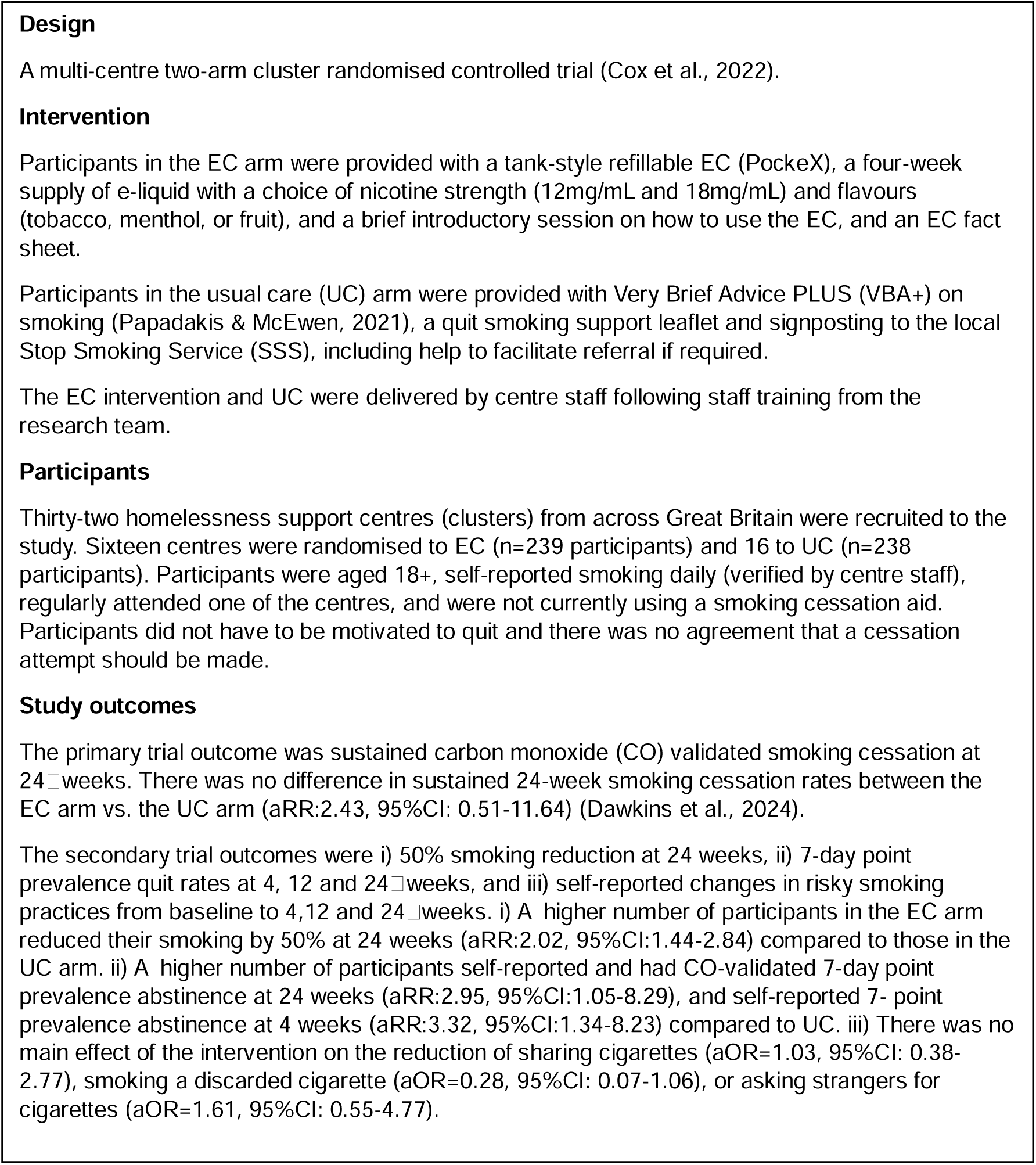
Details of SCeTCH, a cluster randomised control trial

## METHODS

### Design

Using the Medical Research Council framework for developing and evaluating complex interventions (Moore et al., 2015), an embedded mixed method process evaluation was designed around three research questions (RQ). RQ1: How is the EC intervention implemented and how does organisational and geographic context influence implementation? RQ2: What are the mechanisms through which the delivered intervention activities and participant interactions produce change in smoking behaviour? RQ3: If the intervention is effective and cost-effective, what are the facilitators and barriers to roll out across Great Britain? An overview of the process evaluation methods (observations, fidelity checklists, staff training evaluation forms, questions within participant case record forms (CRFs) at baseline and follow-ups, in-depth qualitative interviews), and how they were designed to answer the RQs is outlined in the study protocol (Cox et al., 2022). The trial was registered on the ISRCTN registry (ID:18566874) and Open Science Framework https://osf.io/yhmk9/. Ethical approval for the study was granted by London South Bank University ethics committee (ETH 2021-0176/2122-0130/2122-0142).

This paper addresses RQ2. Quantitative measures placed within CRFs, and interviews with centre staff and participants, were designed to explore hypothesised and emerging mechanisms of change and the mediators of these. Thirteen mediators for changes in smoking behaviour were developed, drawing on the Capability Opportunity Motivation – Behaviour (COM-B) model (Michie et al., 2011), and set out in the SCeTCH logic model (Supplementary figure 1).

### Setting

SCeTCH took place in 32 centres for homelessness across six areas of Great Britain: Scotland (n=6), Wales (n=4), Southwest England (n=2), East England (n=7), Southeast England (n=6) and London (n=7). Centres were a mix of daytime drop-in and residential centres which also offered support for non-residents during daytime hours.

### Sample

Between February 2022 and June 2023, 477 people accessing the 32 centres for homelessness, who reported regularly smoking were recruited to SCeTCH. All intervention participants (n=239) were included in the quantitative sample for this paper. For the qualitative sample, a subsample of 31 participants took part in a process evaluation interview. Interviews were also conducted with 16 staff members. We purposively sampled 8 centres (4 England, 2 Wales, 2 Scotland) within which to conduct the interviews, according to factors such as centre size (i.e. number of daily attenders/residents), types of service provision (day centre/residential), staffing and services offered. This allowed for variation in the centres selected. Within each of these 8 centres, and to aim for a varied qualitative sample in terms of vaping/smoking and sociodemographic status, we selected and invited 3-4 intervention participants to take part in an interview, along with 2 staff members. The purposive sample was identified through discussion and agreement among the research team.

### Procedure

All trial participants gave informed written consent to i) take part in the study and ii) be contacted regarding participation in a process evaluation interview. Researchers obtained initial consent face-to-face prior to completion of the baseline CRF. Participants and staff gave separate written consent to take part in an interview.

#### Quantitative measures

Questions were developed around the COM-B and inserted into CRFs (available at https://osf.io/yhmk9/), completed at baseline and 4-, 12-and 24-week follow-up appointments. All CRFs were administered by the researcher. Under the ‘Capability’ theme, participants were asked for their perceptions on: i) EC harm relative to regular cigarettes (less harmful than regular cigarettes, equally harmful, more harmful than regular cigarettes, don’t know); and ii) how helpful EC were in resisting the urge to smoke (ranging from 1 = not at all helpful to 5 = extremely helpful). Under the ‘Opportunity’ theme, participants were asked for their perceptions on: iii) whether they felt encouraged or discouraged to use the EC when at the centre(ranging from 1 = strongly encouraged to 5 = strongly discouraged); iv) support from centre staff and other service users to use the EC, (ranging from 1 = a lot to 4 = not al all); v) acceptability among staff for service users to use ECs (ranging from 1 = acceptable to 5 = unacceptable; vi) the amount of centre service users using ECs (all of them, most of them, about half of them, a few of them, none of them, don’t know); and vii) engagement with risky smoking practices (sharing cigarettes, smoking discarded cigarettes, asking strangers for cigarettes) (not at all, occasionally, regularly, daily). Under the ‘Motivation’ theme, participants were asked about: viii) motivation to quit smoking, using the one item Motivation to Stop Smoking Scale (MTSS) (Hummel et al., 2017), (ranging from 1 = I don’t want to stop smoking to 7 = I really want to stop smoking and intend to do so in the next month); ix) whether ECs can help people stop smoking or reduce how much they smoke, (ranging from 1 = strongly agree to 5 strongly disagree); x) their average weekly spend on tobacco; and xi) their ability to reduce or stop smoking, (yes – definitely, yes – I think so, I’m not sure, no – I don’t think so, no – definitely not). Patient and public involvement (PPI) representatives were consulted on the appropriateness and wording of all questions with the CRFs.

#### Qualitative interviews

Individual in-depth interviews with participants were conducted between participants’ 12-and 24-week follow-up appointments to allow for longer-term development of the hypothesised mechanisms of change. Interviews took place face-to-face in a private space at the centre, were conducted by AF, LM, EW, AV, CM, JL, JB, AE and DM, and lasted on average, 32 minutes. Staff interviews took place between weeks 4 and 8 after the start of the intervention and were conducted by telephone, by researchers not involved in training delivery or data collection at the same centre, to reduce social desirability bias. Staff interviews were conducted by AF, LM and RB, and lasted on average, 36 minutes. All interviews were audio-recorded with participants’ consent. Separate participant and staff topic guides were informed by the study logic model (Supplementary figure 1) and feasibility study findings (Cox et al., 2021) and further developed in consultation with PPI representatives. Participant interviews covered background information and smoking history, experiences of the intervention, social influences and support for using the EC, and motivations for and feelings about, smoking/vaping. Staff interviews covered background and context/centre information, perceptions of the staff training delivered by the local research team, perceptions and experiences of the intervention, and views on the sustainability of the intervention.

### Analysis and reporting

Analysis followed the process evaluation analysis plan set out in the protocol (Cox e al., 2022). We report descriptive statistics for the quantitative evaluation measures at each time point for those participants who provided a response. For some measures no baseline score was recorded. Quantitative analyses were conducted in Stata v18.0. Interviews were transcribed and de-identified for analysis. Analysis was conducted using a Critical Realist approach and included deductive (from the topic guides and hypothesised mediators in the logic model) and inductive (from interviewees’ accounts) approaches. Two initial thematic coding frameworks (one for participant and one for staff data) were developed by EW, LM, and AF, based on reviewing a selection of transcripts. Independently, EW, LM, JL and AF tested the frameworks against a random 10% sample of the transcripts, and then met to discuss and refine these. Transcripts were coded in NVivo12 by LM, DM, EW, AV, JL, BG and CM, and written summaries of the coded data were prepared by LM and BG. Both quantitative and qualitative data were mapped onto the COM-B model (Michie et al., 2011) and the hypothesised mediators for capability, opportunity and motivation. The inductive approach to qualitative analysis allowed for the identification of additional emerging mediators, also mapped onto the COM-B model. For each COM-B component, quantitative and qualitative data were combined at the interpretation and reporting level. The qualitative findings were also summarised into barriers and facilitators. Initial results were drafted by LM and AF and discussed with the wider team until consensus was reached. To indicate the frequency with which themes were provided by interview participants we use “all”, “most”, “many”, “some”, and “a few”. Quotations have been selected to evidence and illustrate key findings. To minimise any chance of identification, participant quotations indicate their vaping/smoking status only. Staff quotations include their role. To minimise any bias that might be introduced by their beliefs, the authors involved in the analysis reflected on the potential impacts that their shared identity as white female researchers working in tobacco harm reduction might have on the findings.

## RESULTS

The characteristics of the quantitative participant sample and the subsample and staff who took part in an interview are reported in Table 1. The qualitative sample broadly mirrored the wider quantitative sample in terms of age, gender, ethnicity and education.

**Table 1.** Participant characteristics.

|  |  |  |
| --- | --- | --- |
| **Intervention participants** |  | **n= 239** |
| Age (y), mean (SD) |  | 42.1 (11.0) |
| Gender N (%) |  |  |
| Male |  | 193 (80.8) |
| Female |  | 44 (18.4) |
| Non-binary |  | 1 (0.4) |
| Transwoman |  | 1 (0.4) |
| Ethnicity, White N (%) |  | 198 (82.9) |
| Education N (%) |  |  |
| Stopped before GCSE or equivalent |  | 93 (38.9) |
| GCSE or equivalent |  | 88 (36.8) |
| A-level or equivalent |  | 42 (17.6) |
| University degree |  | 13 (5.4) |
| University Postgraduate degree |  | 3 (1.3) |
| **Intervention participants – interview subsample** |  | **n= 31** |
| Age |  |  |
| 18-30 |  | 3 |
| 31-40 |  | 8 |
| 41-50 |  | 6 |
| 51-60 |  | 11 |
| 61-70 |  | 2 |
| Prefer not to say |  | 1 |
| Gender |  |  |
| Male |  | 23 |
| Female |  | 7 |
| Non-binary |  | 1 |
| Ethnicity |  |  |
| White |  | 28 |
| Black |  | 2 |
| Mixed |  | 1 |
| Education |  |  |
| Stopped before GCSEs or equivalent |  | 10 |
| GCSEs or equivalent |  | 11 |
| College |  | 6 |
| Ethnicity |  |  |
| White |  | 28 |
| Black |  | 2 |
| Mixed |  | 1 |
| Education |  |  |
| Stopped before GCSEs or equivalent |  | 10 |
| GCSEs or equivalent |  | 11 |
| College |  | 6 |
| University |  | 4 |
| 12-week follow-up smoking/vaping status |  |  |
| Exclusively vaping |  | 2 |
| Dual using |  | 20 |
| Non-smoking/-vaping |  | 1 |
| Exclusively smoking |  | 8 |
| **Staff** |  | **n = 16** |
| Gender |  |  |
| Male |  | 7 |
| Female |  | 9 |
| Staff role |  |  |
| Managerial |  | 8 |
| Support worker |  | 8 |

Quantitative and qualitative findings for hypothesised and emerging mediators are set out under the COM-B headings Capacity, Opportunity and Motivation. Table 2 presents an overview of the quantitative results. Table 3 summarises the qualitative findings into barriers and facilitators to smoking behaviour change.

**Table 2.**
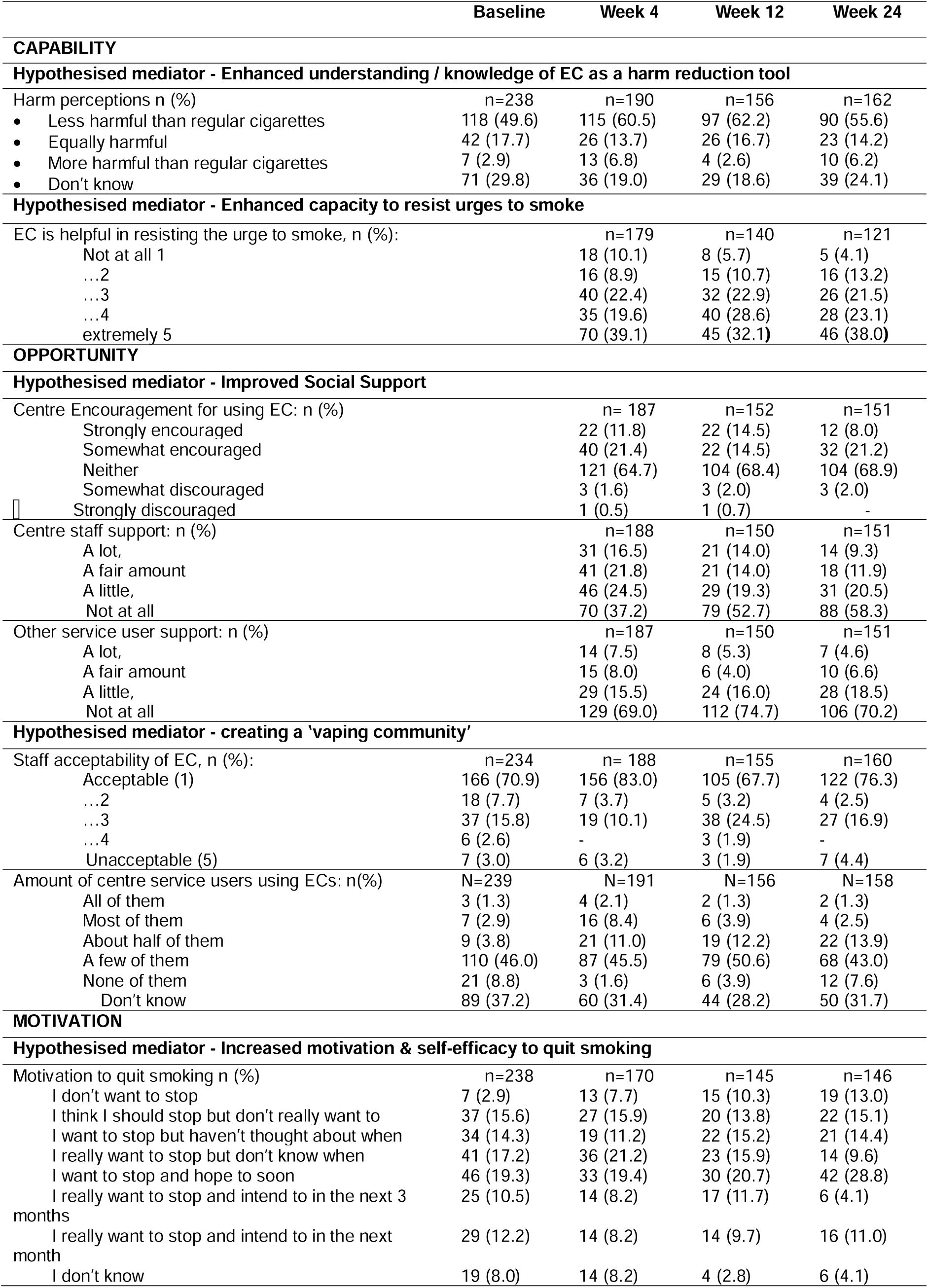
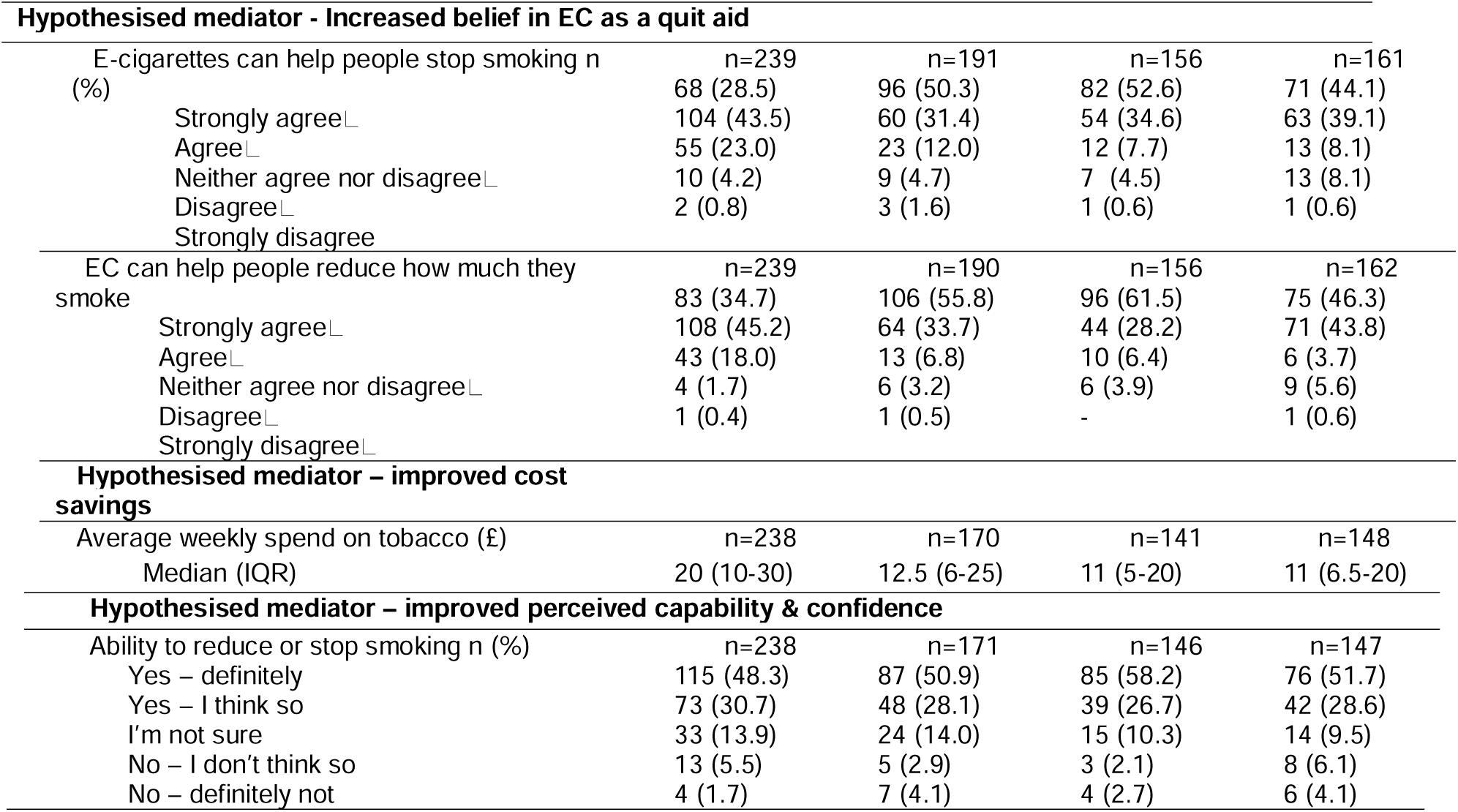
Quantitative CRF measures, mapped to the COM-B Framework.

**Table 3.**
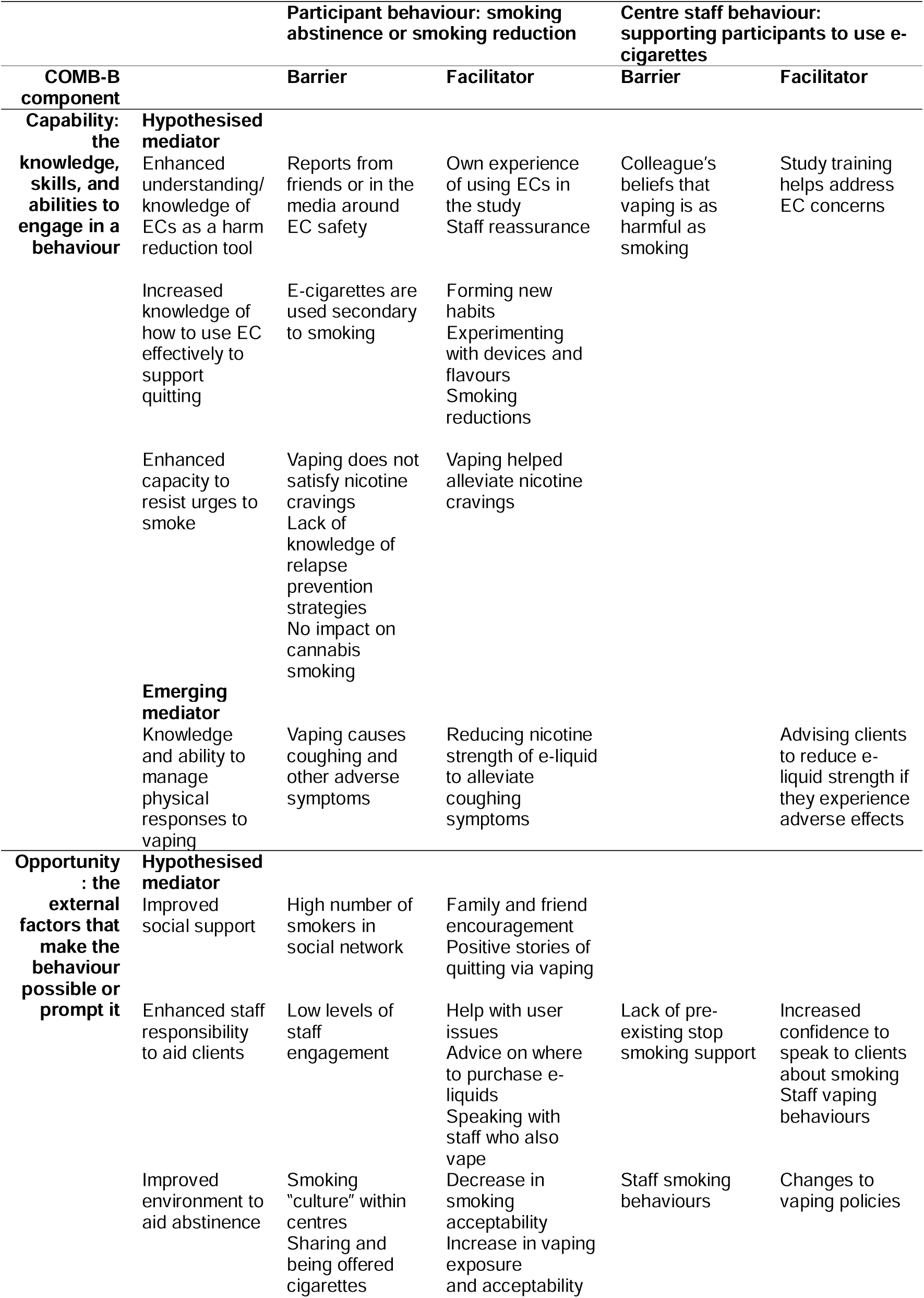
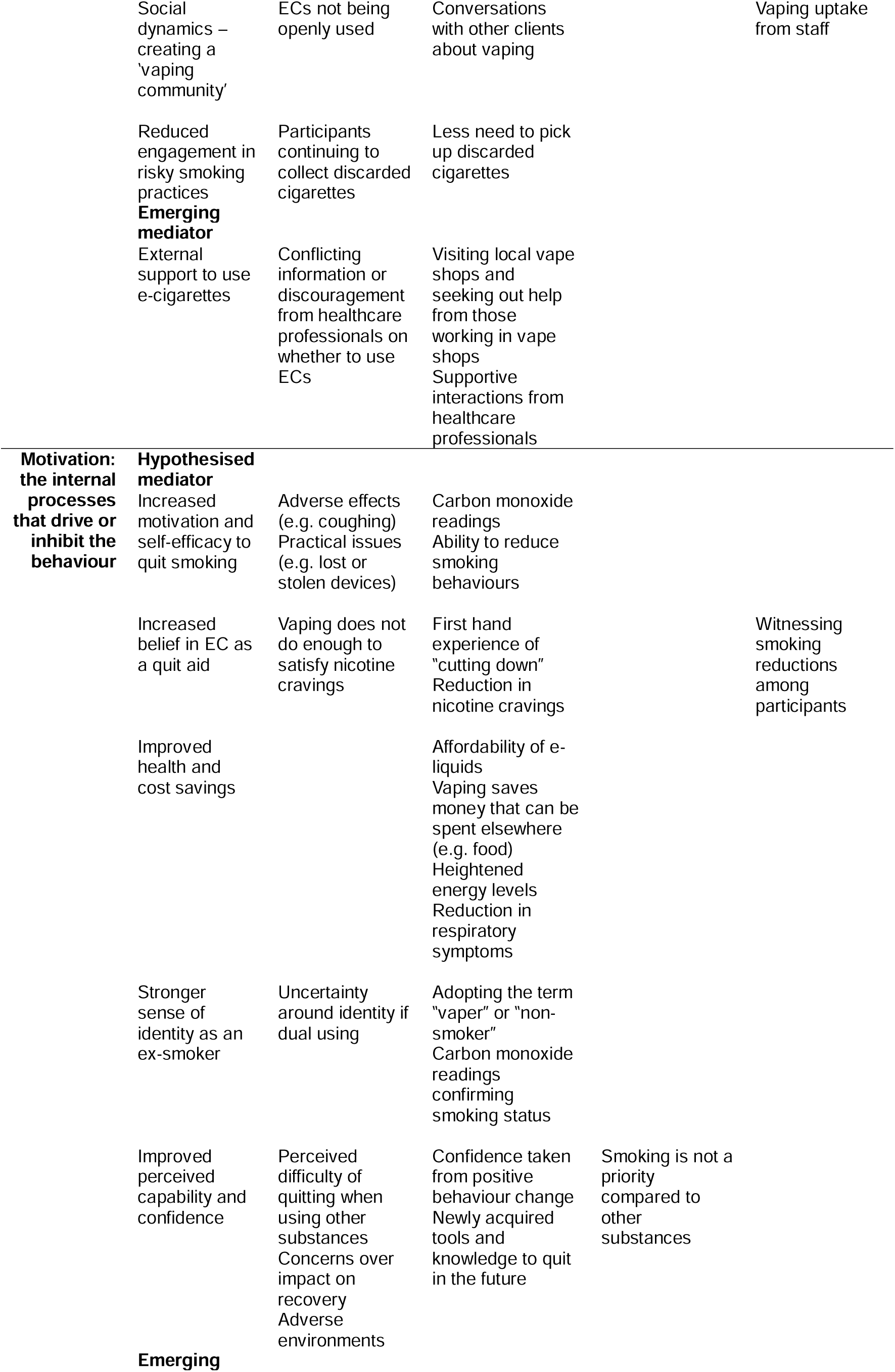
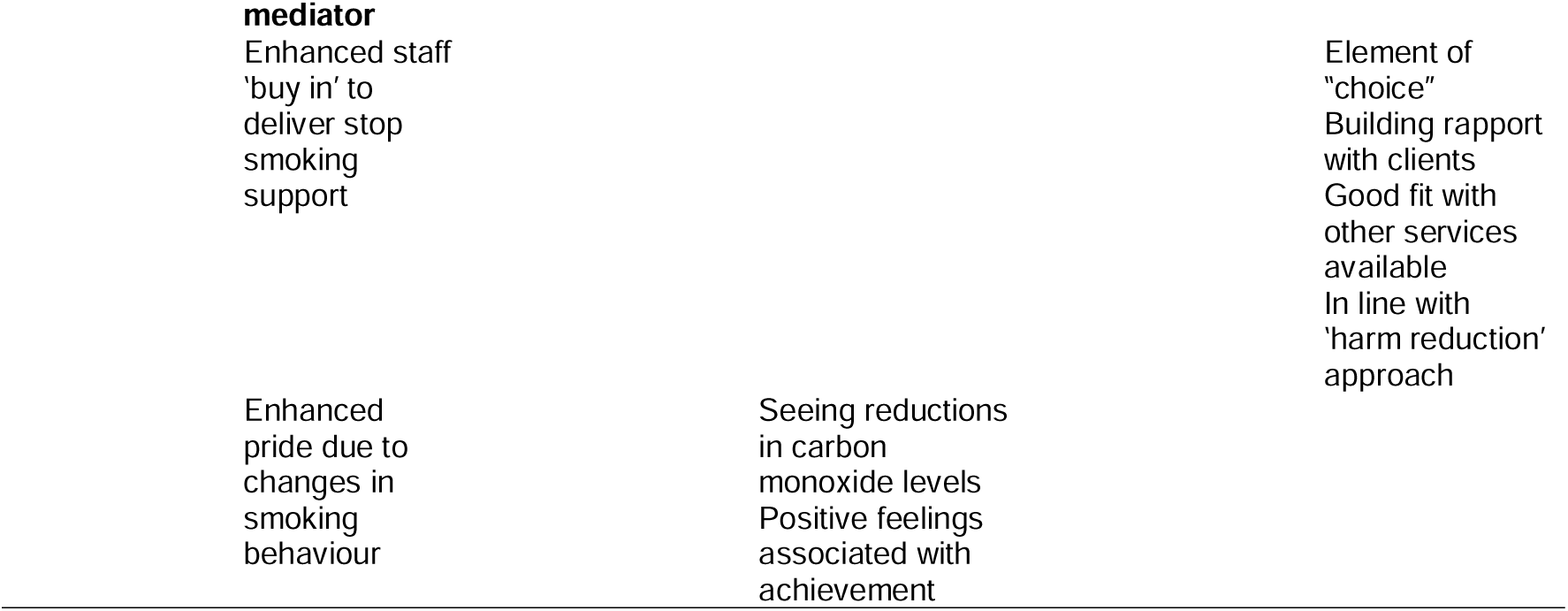
Qualitative summary of hypothesised and emerging mediators, categorised as barriers and facilitators to changes in smoking behaviour, and mapped to the COM-B framework.

### Mechanism of change: Capability

#### Hypothesised mediator - Enhanced understanding/knowledge of e-cigarettes as a harm reduction tool

Most participants reported they perceived EC as less harmful than regular cigarettes at all timepoints (Table 2). At baseline 49.6% responders reported perceiving EC as less harmful. Among those responding at 24 weeks, this had increased to 55.6%.

This is reflected within the qualitative data, with some describing how their positive experiences with the intervention helped strengthen beliefs that EC could be used as a harm reduction tool. This was despite having heard stories in the media or through friends that contributed to initial doubts around EC safety (Table 3).

> *“When [EC] first come out, yes, I would be a little apprehensive because nobody’s done a proper study on them yet…but since having them, I’d say they’re less harmful than cigarettes.”* (Dual using)Some participants, however, felt there was no difference between the harm caused by smoking and vaping.

> *“I can’t see that there’s any difference…you’re still putting something carcinogenic through your throat, you’re inhaling it in your mouth…it’s going into your blood vessels.”* (Dual using)The qualitative data demonstrated that for most staff, the training delivered by the research team was pivotal in enhancing their knowledge of EC as a harm reduction tool, enabling them to discuss the benefits of EC for quitting smoking and confidently recommend EC to participants.

> *“We were armed with the knowledge of being able to actually challenge those notions that the service users may have had for a very long time and stigma against using an electronic vape.”* (Support Worker)Lack of understanding around EC harm was still evident within some centres. One staff member reported pushback when they proposed an indoor vaping policy because the centre manager, who had not attended the training, believed vaping was “*just as bad as smoking*” (Support worker).

#### Hypothesised mediator - Increased knowledge of how to use e-cigarettes effectively to support quitting

Most interview participants demonstrated high levels of capability to use their EC to support quitting. They demonstrated practical knowledge on using the device and recalled forming new habits to reduce their smoking, for instance, vaping in between favourite cigarettes, vaping at specific times of the day, or swapping certain cigarettes in favour of vaping.

> *“I started taking my e-cig with me instead of tobacco, and I started smoking my e-cig before training and after training, and it felt well better.”* (Dual using)Participants demonstrated increased capabilities by experimenting with different types of e-liquid. For some, this was within the scope of the study (changing nicotine strength or switching flavour). Others enjoyed testing out a range of e-liquids they had purchased.

Participants also reported experimenting with other EC devices, both refillable and disposable, until they found one more suited to their needs.

> *“I had [the study e-cigarette] for a while, then realised I wasn’t using it. I then bought something else…It was like a salt one and I thought, ‘oh, that is more like it…it has just got a better flavour…and it just feels better…’ that was the one that clicked. I had to find the right kind of vape.”* (Non-smoking)The majority of interview participants were dual using. Some described vaping as secondary to smoking. For instance, keeping their EC on standby or as a fallback option. Others described vaping as superior to smoking but carried cigarettes “*just in case*” (Dual using) or limited smoking to a few cigarettes they deemed more important (e.g. first one in the morning). Many participants described a reduction in smoking since receiving their EC at the start of the study.

> *“I don’t smoke 20-odd fags a day now…I smoke maybe three…if I didn’t have the e-cig, I think I’d smoke more.”* (Dual using)This idea of cutting down as opposed to quitting was also widely reported by staff.

> *“We see clients that are buzzing…they’re showing me their vapes and how much they’ve cut down, and you get that energy from it, yeah, it’s nice to see that type of thing.”* (Support worker)

#### Hypothesised mediator - Enhanced capacity to resist urges to smoke

Most participants (50% or more), across all time points, positively reported that the EC was helpful in resisting the urge to smoke although there was some variation (Table 2).

The qualitative data supports this mixed view. Some participants described how vaping had helped alleviate their nicotine cravings and for one participant, this meant they had “*no desire to go back to a cigarette*” (Exclusively vaping). Others reported that they were unable to satisfy their urge for nicotine through vaping in the same way they could through smoking, which impacted their ability to stay smoke free.

> *“I need a nicotine hit…don’t get me wrong, the vape can do it for so long, but I still need the fags as well.”* (Dual using)Dissatisfaction of vaping compared to smoking was one of the main reasons cited by participants for relapsing to smoking. Despite high levels of motivation to stay quit, some participants discussed how events in their lives had impacted their ability to stay smoke free. Many were dealing with multiple issues at the same time.

> *“…the next day I just collapsed…not because I didn’t want to stop smoking, it was because a lot of stuff was going wrong…I just couldn’t cope with it no more.”* (Exclusively smoking)Those who smoked tobacco and cannabis reported that while vaping reduced their nicotine cravings and thus, the number of cigarettes smoked, they continued to smoke tobacco with cannabis at the same rate as they had prior to receiving their EC.

#### Emerging ‘Capability’ mediator – Knowledge and ability to manage physical responses to vaping

Within the interviews, almost half the sample reported experiencing coughing when using their EC. They described it as a “*catch in the back of your throat*” (Dual using) type cough, or used adjectives such as too “strong”, “powerful” or “harsh” when talking about the device or e-liquid. Staff members also picked up on participants coughing when first using the device and offered insights into whether participants were able to manage this.

> *“The first initial [attempts to use], a lot of them were coughing, because it’s a different technique to smoking…that went away as they got used to it. And some even changed the strength of their liquid because of it too.”* (Manager)Some participants explained how they were able to alleviate their symptoms and continue vaping after speaking with researchers, staff or family members who had advised them to switch to a lower nicotine strength e-liquid.

> *“Initially, when I tried these [e-liquids], they’d make me cough…my brother said to me, ‘it’s because these are so high in nicotine’…I explained that to [researcher] and [she] put me on a lower dose, not the 2000 or whatever, she put me on the 1200. And I don’t cough half [as much]…if I puff on those…” (Dual using)*However, a few participants were unable to manage the coughing they experienced when using the EC, which led to them stop vaping altogether.

> *“I did try [vaping]…but the coughing was really, really bad…I was just like, ‘oh, I can’t do that anymore…I need to give that up’.”* (Exclusively smoking)

### Mechanism of change: Opportunity

#### Hypothesised mediator - Improved social support

Most participants, at all timepoints, reported feeling neither encouraged nor discouraged to use their EC when at the centre (Table 2). By week 24, the majority (58.3%) of respondents reported that they felt not at all supported by staff to use their EC while at week 4 this proportion was 37.2%. This is also reflected in the proportion of those reporting being supported by staff to use their EC decreasing from 38.3% at week 4 to 21.2% at 24 weeks.

Although the hypothesised increase in social support in intervention sites was not observed, many interview participants reported high levels of support to quit smoking from people beyond the centre, such as their friends and family. Participants referred to people close to them who also vaped, and this provided opportunities to hear positive experiences of quitting using EC and to discuss or try different products.

> *“[My brother] has offered to take me to the vape shop…to buy me a new vape and [e-liquid] and pay for everything.”* (Dual using)Some recalled feeling encouraged by their friends and family to use their EC and described how supportive these individuals had been when they heard they had cut down or quit smoking.

> *“I had my daughter, and my son-in-law, they were really supportive…there was loads of people saying…‘if you stick to [vaping]… you would be doing great.’”* (Dual using)However, many participants reported high levels of smoking exposure across their social networks. This presented more opportunity to smoke rather than vape.

*“I haven’t got a big circle of friends but the people I do know, they do smoke. So it’s always encouraged to have a cigarette…” (Dual using)*

#### Hypothesised mediator - Enhanced staff responsibility to aid abstinence

As noted above, perceived support from staff decreased over the course of the study. Qualitative data on whether staff took responsibility to support participants to quit smoking were mixed. Some participants reported help with user issues and advice on where to buy affordable e-liquids. However, there were accounts from others who did not feel supported to use their EC or that it was at their own discretion whether they used the device. There was a sense among some participants that staff could have done more to support those taking part.

> *“I think the staff could put a bit more effort into engaging with…the residents…I think they would find that the residents would be more eager than they thought.” (Exclusively smoking)*Staff noted that stop smoking support was mostly non-existent at their centres prior to the intervention. They reported that taking part in the study meant they felt more confident to have conversations around smoking cessation.

> *“Smoking hasn’t really been something we have had a major focus although it is something that we really wanted to do…what the study has allowed us to do is look at things in a slightly different way…it has actually allowed us to go, ‘well, no, let’s actually deal with what the person wants to deal with and bring in smoking.’” (Manager)*In some instances during the trial, staff had been motivated to change their own smoking behaviour and tried to quit via vaping. This was perceived to aid staff engagement with the trial and their support and encouragement for participants.

> *“[My keyworker] is actually on a vape as well…he’s been smoking for quite a few years…but he’s enticing us to stop a lot more. He’s been very supportive.” (Dual using)*

#### Hypothesised mediator - Improved environment to aid abstinence

Many interview participants described a smoking culture within centres that provided opportunities for clients to engage in smoking and did little to aid abstinence. One participant described how “*cigarettes are not frowned upon in the centre*” (Exclusively vaping) and there were few incentives for encouraging clients to quit. Despite their well-meaning intentions, some staff members facilitated smoking for clients.

> *“I keep a pack of cigarettes here if somebody is stressed out…sometimes a cigarette will help them calm down a bit, which I don’t mind then doing now and then.” (Support worker)*Sharing and being offered cigarettes was widely reported across the centres, which impacted participants ability to stay smoke free.

> *“There is one friend [at the centre]‘ she’ll just pass me the package and I’ll just have one…it’s not in a bad way, it’s a decent way. But this is what drives me mad, because I’m thinking I want to stop but I’m so easily led by just a cigarette on offer…” (Dual using)*Staff similarly described the centres as high smoking environments with no separation between vaping and smoking areas. When staff were asked if they reviewed any of their existing smoking or vaping policies due to taking part in the trial, only one centre had implemented a change to their policy.

Many staff discussed stop smoking support in comparison with other forms of substance use support available at the centre. There was a perception that supporting clients to abstain from drug use or help them with their alcohol problems took precedence over smoking. One staff member explained that in terms of “*the hierarchy of harm*” (Support worker) there were more pressing issues than smoking that staff members tended to focus their attention towards.

> *“We work a lot to try and reduce kind of people’s class A drug use and alcohol use…that has a significant effect on somebody’s life and future and plans…that’s more of a priority.” (Support worker)*Beyond the centre, there were more opportunities that aided quitting. Some participants had noticed a decrease in smoking acceptability and explained that they preferred vaping socially as “*there’s not so much stigma around it*” (Dual using). Some also highlighted the benefits of being able to use their EC in environments where smoking was less acceptable and noted the positive influence of vaping exposure.

> *“The more I see people vaping, the more I’m encouraged to vape and I see…lots of people outside vaping rather than the cigarette.” (Dual using)*

### Hypothesised mediator - Social dynamics – creating a ‘vaping community’

Most participants, across all timepoints, perceived that if they used EC it would be acceptable to staff (Table 2). The proportion of responders who perceived staff acceptability was similar at baseline and 24 weeks (78.6% and 78.8% respectively). Around one third of responders, at all timepoints, did not know how many other service users at their centre used EC.

Reflecting the strong smoking norms described above, while some participants discussed seeing others at the centre vaping, most recalled seeing little evidence of a vaping community forming within the centre, for example seeing others vaping, or vaping together.

> *“I don’t know how many you’ve gave out, but I haven’t seen many people using them.” (Dual using)*Despite this, both staff and participants reported how taking part in the trial had triggered conversations around vaping or had an influence on other clients who were now interested in trying vaping after seeing some intervention participants “*reaping the benefits*” (Support worker).

> *“… [the study e-cigarette] was very popular among my friends…‘oh this is amazing, oh I’ve never seen one like this before, where did you get this’, et cetera. So many compliments…” (Dual using)*While many participants felt that their vaping did not have an influence on others, there were a few examples where staff had either started vaping or considered vaping since delivering the intervention to participants.

> *“I was sceptical in relation to the e-cigs. It wasn’t something I was sold on. but after this research or during it I actually tried one and I was like okay, and it’s actually helped me reduce my smoking.” (Manager)*

#### Hypothesised mediator - Reduced engagement in risky smoking practices

Quantitative findings on participants’ reporting of risky smoking practices are presented elsewhere.^23^ There was a reduction over time in the proportion of responders who reported smoking discarded cigarettes but not sharing cigarettes or asking strangers for cigarettes.

Within the qualitative data, there were few narratives from participants and staff around whether the intervention had reduced participants’ engagement in risky smoking practices. While one participant stated that they hadn’t picked up discarded cigarettes for over a month since starting the study, another reported seeing participants continuing to engage in risky smoking practices.

> *“…some [service users] that I do know of on the trial are still walking out to that bin out there and get all the [discarded cigarettes] out them and make themselves roll-ups…” (Dual using)*

#### Emerging mediator – External support to use e-cigarettes

Interview participants detailed other forms of opportunities for support they had received beyond the hypothesised mediators. Some participants had visited vape shops during the study and recalled “friendly” interactions and “helpful” advice from staff around which device to use and e-liquid to buy.

*“I’ve got a few different ones I go to…some of them are very helpful with information, tell you…what sort of vape is good for that…what vape is good for your CBD…” (Dual usin*

Some participants also reported supportive interactions with healthcare professionals away from the centre who encouraged them to quit smoking and try vaping instead. However, a few participants described a lack of cessation support or conflicting information on vaping when speaking with their General Practitioner about their smoking.

> *“[My doctor] went through my medical history and said that I shouldn’t really be smoking at all. But when I told him about vapes…he would rather I didn’t have them either.” (Dual using)*

### Mechanisms of change: Motivation

#### Hypothesised mediator - Increased motivation and self-efficacy to quit smoking

Most participants, across all timepoints, reported that they wanted to stop smoking, although there was some variation in responses between follow-up timepoints (Table 2).

Some interview participants, who reported that their motivation to quit gradually increased as the study progressed, described the positive effect that the intervention had on strengthening their future quitting beliefs. The inclusion of exhaled carbon monoxide readings, taken at baseline and each follow-up appointment, were particularly beneficial at enhancing motivation.

*“…using all of the breath tests, you see a benefit. Which you don’t see normally…it’s showing a reality…you can see it going from maybe 15 down to 11…you can see it as you’re progressing. (Dual using)*

Other participants described their motivation levels wavering as the study continued and credited external circumstances and adverse effects, such as coughing, or practical issues, such as lost or broken devices, as contributing factors. For those who had been unsuccessful in quitting, many remained hopeful that they would continue to use their EC or return to vaping in the future once stressful life events had settled down.

Overall, self-efficacy to quit appeared high among most participants. Their beliefs were bolstered by their ability to reduce the amount of tobacco or cigarettes smoked since receiving their EC.

> *“…[cutting down] has helped…it makes me think, ‘if I wanted to stop smoking, then I could go that little bit further…that I could push myself.’’’ (Dual using)”*

#### Hypothesised mediator - Increased belief in e-cigarettes as a quit aid

Most participants, at all timepoints, agreed or strongly agreed that EC can help people stop smoking or can help people reduce how much they smoked (Table 2). At baseline, 72% of responders reported that EC can help people to stop smoking and at 24 weeks this proportion had increased to 83.2%. At baseline, 79.9% of responders believed that EC can help people reduce how much they smoke and at 24 weeks this had increased to 90.1%.

Within the qualitative data, while a minority of participants did not believe in the efficacy of using EC as a quit aid, many drew on their own experiences of cutting down during the study, which supported the belief that EC “do work” and made them more motivated to use EC to stop or reduce smoking.

> *“If I didn’t have the e-cig, I think I’d smoke a lot more cigarettes…I would smoke a cigarette not just in the morning, dinner time and teatime…I think I would smoke all day…” (Dual using)*Staff also reported that taking part in the study helped to tackle their own scepticisms around the efficacy of EC and made them more motivated to use EC after witnessing smoking reductions among participants or trying vaping themselves.

> *“I was sceptical in relation to the e-cigs. It wasn’t something I was sold on…but after this research or during it I actually tried one…and it’s actually helped me reduce my smoking.” (Manager)*

#### Hypothesised mediator - Improved health and cost savings

Overall, participants reported a reduction in their weekly spend on tobacco between baseline and 24 weeks. At baseline, the median weekly spend reported was £20, for those completing the CRF at 4 weeks it was £12.50, and at both 12 and 24 weeks, this was £11 (Table 2).

Within the interviews, cost saving was the most pertinent benefit that participants reported since receiving the EC intervention. They described how vaping had reduced the amount of tobacco they purchased, and access to e-liquids was at a price that was “*bloody cheap compared to smoking*” (Dual using). Health benefits were also noted. These ranged from improvements in physical health including asthmatic symptoms to improvements in energy levels and overall mental health.

> *“It totally helped me…I didn’t feel lethargic, I didn’t feel fatigue…The cigarettes had caused fatigue, depression…laziness, lack of motivation. But as soon as I started vaping, I didn’t get any of that.” (Exclusively vaping)*Staff members also discussed the financial benefits of switching from smoking to vaping. They felt this was an important motivator for their clients as it allowed them to reallocate the money saved towards other aspects of their lives. There was a noted interaction between this mediator and that of increased motivation:

> *“The free e-cigarette and the follow up vouchers. That was the motivation [for taking part]. Then I think the benefits were then noted by people, that they’re not spending as much money, this isn’t as harmful, this flavour lasts a week while a packet of cigarettes lasts a day, hey, let me work with this.” (Manager)*

#### Hypothesised mediator - Stronger sense of identity as an ex-smoker

Some interview participants described changes to their identities around smoking as their EC use increased or smoking decreased and used terms like “non-smoker” or “vaper” to describe their smoking and vaping status. One participant registered a reading of four on the carbon monoxide breathalyser and was delighted they were classed as a “non-smoker”, while another who had recently quit felt that it was too soon to describe themselves as a “non-smoker”:

> *“It’s early days yet…a fortnight ain’t that long…if it was a few months, I’d say non-smoker, but…I’ve just given up.” (Non-smoking)*Participants who had reduced their smoking or had tried and failed to quit described themselves as a mix of the two. For instance, as “half and half” or “in the middle” between a ‘smoker’ and a ‘vaper’. The term ‘dual user’ was absent from participant’s accounts. One participant highlighted the difficulties in determining their identity now that they were vaping as well as smoking.

> *“…it’s awkward…I’m still a smoker and I’m nearly an e-vaper. But I’m not quite yet…as long as I’m buying tobacco, I think I’m always going to be a smoker…” (Dual using)*

#### Hypothesised mediator - Improved perceived capability and confidence

When asked whether they felt they had the ability to reduce or stop smoking, most participants, at each timepoint, answered ‘yes, definitely’, or ‘yes, I think so’. The proportion of responders answering positively was similar at baseline (79%) and at 24 weeks (80.3%) (Table 2).

Within the interviews, many participants reported how taking part in the trial had increased their confidence with vaping and enhanced their motivation as they were equipped with the tools and knowledge to make a quit attempt in the future.

> *“When I had [the e-cigarette], it was helping…I’ll get myself another one and try and use my oil, since the oil’s just still sitting there.” (Dual using)*Drawing back to one of the opportunity mediators in which some staff perceived that there were more pressing issues that may have hindered the centre’s environment to aid abstinence, staff also highlighted the challenges of offering stop smoking support to clients with complex substance needs. This was echoed by the participants who felt it would be “trickier” for them to quit smoking if they were using other substances or were afraid it might impact their recovery.

> *“…my life is staying sober…it’s not even one day at a time, it’s one hour at a time…so at the moment, I have no plans to stop smoking…my proclivities are my proclivities but it keeps me sober for now…” (Dual using)*Participants who didn’t feel capable of quitting also explained why it wasn’t “the right time” in their lives to make a quit attempt as adverse environments or stressful circumstances meant they were “just not there yet”.

> *“…to put myself through that, sat there…fidgeting, looking at the telly…thinking, I want a fag, it might make me more stressed. And it might be worse for me…I think once I’m settled in my place…” (Dual using)*

#### Emerging mediator - Enhanced staff ‘buy in’ to deliver stop smoking support

Interviews with staff highlighted that their motivation to deliver stop smoking support was driven by direct and indirect components of being part of the trial. Staff members spoke about the element of choice as an important factor of the study that resonated and they liked that EC provision had given their clients “access to the same things” as everyone else. They also liked, that through the study, they were able to build rapport with clients. Connecting with clients through the study provided opportunities for offering additional support.

> *“It gave people an excuse to come and see me, and when you’re talking to them about that, you can talk to them about something else.” (Support Worker)*Staff also explained being able to offer stop smoking support was a good fit with the other services that they provided, such as health check-ups or drug and alcohol support, thus enhancing the overall quality of care that clients could receive at the centre.

> *“We have the dental nurse in…we have the sexual health nurse in…I don’t see why…stopping smoking should be out of that equation the same as brushing your teeth…or your physical health as well.” (Support Worker)*Similarly, there were staff members who felt that the intervention complimented the harm reduction approach to substance use that their centre embraced and that their clients were already familiar with.

> *“Harm reduction…it’s not a…foreign concept, and not a shameful thing…if you reduce a bit, then there’s loads of positives to gain from that, and I suppose we’re equally the environment that supports that…” (Manager)*

#### Emerging mediator - Enhanced pride due to changes in smoking behaviour

Positive mood was enhanced by the feelings that were evoked after cutting down the amount of tobacco they were smoking. Some participants described how “happy” or “pleased” they were that they had been able to make positive changes to their smoking behaviour, while others elaborated on the pride they felt seeing a decrease in their carbon monoxide readings.

> *“Blowing on that machine each time, it’s dropping, and that’s like, ho, whoa…from 30 to 16, that’s a big drop. I’m proud of myself because of that.” (Dual using)*Among those who had cut down, these positive emotional responses were also key in strengthening their intentions and beliefs in their ability to quit smoking in the future.

> *“I’m pleased I done it…I feel as if I’ve achieved something…it’s gave me a wee bit of confidence that, if I wanted to stop…then I could go that little bit further…that I could push myself.” (Dual using)*

## DISCUSSION

This study explored the mechanisms which served as barriers and facilitators to smoking reduction and short-term, but not sustained, abstinence from an EC intervention delivered at homelessness support centres. Using the COM-B model, we explored hypothesised and emerging mediators through Capability, Opportunity and Motivation themes for smoking behaviour change.

Participants demonstrated high capability to use EC. We found good levels of knowledge of EC as a harm reduction tool, enhanced for participants through positive intervention experiences, and for staff through training. Although quantitative data indicated a majority belief that EC were helpful in resisting the urge to smoke, interviews highlighted some dissatisfaction with vaping compared to smoking, and many participants were dual using.

Some had to manage coughing and experienced the EC and e-liquid as too ‘strong’ or ‘harsh’. These factors, coupled with stressful life events and low ability to abstain from smoking, explain how many participants could not completely switch to vaping.

Opportunity for behaviour change was strengthened by support and encouragement to vape, particularly from friends and family but also vape shop staff and health professionals beyond the centre. Although quantitative data showed high levels of perceived acceptability to vape at the centre, there were mixed qualitative accounts of staff supporting or encouraging participants to vape and little evidence of a vaping community forming at centres. However, both participants and staff said the intervention had triggered discussions around vaping, and some staff who had been motivated to switch to vaping because of the trial, were more engaged with encouraging participants to quit. Opportunity for smoking abstinence was hindered by a smoking culture where tobacco was commonly offered or shared.

Motivation was enhanced by strong beliefs in EC as a quit aid, positive emotional responses from being able to cut down through vaping and observing financial benefits of vaping compared with smoking. Confidence and self-efficacy to quit were also driven by cutting down, and enhanced quitting beliefs for the future. Motivation was negatively impacted by coughing, device issues and challenging personal circumstances. While there was staff ‘buy-in’ to offer stop smoking support, supporting clients with their drug and alcohol use or mental health concerns remained more of a priority.

Other studies have similarly highlighted the challenge of sustained quitting among disadvantaged populations (Gilbody et al, 2019; Vijayaraghavan et al., 2020). Although there is evidence that EC with nicotine may increase quit rates in the general population compared to nicotine replacement therapy, EC without nicotine, and behavioural or no support (Lindson et al., 2024), vaping satisfaction plays a key role in quitting (Yong et al, 2019; Dawkins et al., 2015). It is difficult to know whether the dissatisfaction of vaping reported by some of our participants is due to high levels of tobacco/nicotine dependence which only smoking can satisfy, or the belief in the need for cigarettes for certain situations, i.e. times of acute stress, which affirms tobacco dependence (Notley et al. 2021). The coughing experienced by some of our participants was attributed to high nicotine strength e-liquid, yet dissatisfaction suggests nicotine levels were too low (Voos et al., 2019). While some were able to manage this, it led others to give up trying vaping. Previous work on relapse has found negative experience to be a key reason people give up trying to vape and relapse to tobacco (Notley et al. 2019). Building greater knowledge around puffing technique, i.e. among those delivering the intervention, or encouraging experimentation to find a more satisfying device or tolerated e-liquid, i.e. nicotine salt formulations (O’Connell et al, 2019), may help participants’ capability to make a complete switch to vaping. Many of our participants who were dual using retained their smoker identity, which has also been implicated in continued smoking (Notley & Collins, 2018).

Our findings show that staff felt empowered to have smoking-related discussions with service users because of the training they received. Yet participants perceived lack of support by staff. This may be due to a trial fatigue effect, where initial enthusiasm to support the intervention waned over time. Top-up training sessions for staff may help to sustain ‘buy-in’ and reach staff who had not had an opportunity to attend previous training, or who may be sceptical of the intervention or efforts to help people stop smoking. The latter likely reflects practitioner focus on immediate and acute harm of drug and alcohol use or mental health concerns, rather than longer-term harm of tobacco smoking (Taylor et al., 2012). A shift in mindset around the harm of substances is required if staff are to accept the importance of smoking cessation.

Our findings support previous studies which highlight the importance of social and environmental factors when attempting to stop smoking (Pratt et al., 2019). Participants highlighted a lack of skills and tools to enable them to abstain from smoking during times of stress and centre environments which were conducive to smoking. In our feasibility study we observed a culture which shifted from smoking to vaping at one centre, along with higher switching rates (Cox et al., 2021). This was likely due to larger numbers of participants taking part and several staff simultaneously trying vaping. We did not observe such a shift within this study, likely due to smaller numbers participating at each centre. Given perceived financial savings of vaping compared with smoking, it will be important to monitor the potential impact of future taxation (HM Treasury, 2024). If vaping does not remain cheaper than smoking, it may lower motivation to use EC in a future quit attempt, and disproportionately affect disadvantaged populations, potentially widening health inequalities.

### Strengths and limitations

To our knowledge, this is the first study to explore the mechanisms through which an EC intervention produced changes in smoking behaviour among people accessing centres for homelessness. A strength of the study is the use of a theoretically based framework and logic model which informed the quantitative measures and qualitative topic guides. We found the COM-B a useful way to systematically structure data collection and analysis, and it allowed exploration of both hypothesized and emerging (inductively derived) mediators.

Through our analysis some emotional, interpersonal and environmental factors emerged, however, these important influences on behaviour may be more comprehensively explored within a more detailed framework such as the Theoretical Domains Framework or a social-ecological framework (Michie et al, 2005; Cane et al., 2012; Sallis et al, 2008).

The study has some limitations. The CRF measures captured participants’ perceptions of EC at each timepoint, however, the number of participants completing CRFs reduced from baseline, with those remaining possibly having more positive views. This reflects the trial retention rate, which compared favourably to other trials among similar populations (Vijayaraghavan et al., 2020). Despite loss to follow-up, however, the quantitative findings largely reflected the qualitative insights. While the CRF measures involved PPI input there may have been variation in participants’ interpretations of the questions. While the quantitative data are limited in what they tell us about what influences smoking behaviour change, a key strength of the study is the use of qualitative data which provides rich insight into participants’ experiences. Some participants may have given socially desirable responses during CRF completion and interviews. To reduce this possibility, the researchers were careful to use open and balanced questioning and staff interviews were undertaken by researchers not known to them to facilitate open disclosure of their thoughts and experiences. Finally, the qualitative findings may not be generalisable to the trial sample or other people accessing centres for homelessness. Our sample also lacks ethnic diversity.

## Conclusion

These findings show that among people accessing centres for homelessness, achieving smoking abstinence remains challenging. The delivery of an EC intervention led to smoking reduction and short-term abstinence. Enablers included appreciation of EC as a harm reduction tool and positive emotional responses to experiences of cutting down. Barriers to sustained abstinence included lack of satisfaction with EC compared to smoking, a strong smoking culture at centres, and staff deprioritising tobacco-related harm. People experiencing homelessness may require more intensive support and a variety of approaches to help them stop smoking, particularly those which address the psychosocial factors which hinder smoking abstinence. Future research should focus on how this can be achieved.

## Ethical approval

Ethical approval for the study was granted by London South Bank University ethics committee (ETH 2021-0176/2122-0130/2122-0142).

## Supporting information

Supplementary figure 1 SCeTCH trial logic model

## Data Availability

All data available are contained in the manuscript.

## Acknowledgements

The authors would like to thank: members of the Trial Steering Committee; Data Monitoring and Ethics Committee; our PPI group; all centre staff and participants who contributed to the trial; and Dr Laura Reid for proof reading the manuscript. This study was funded by the National Institute for Public Health (NIHR132158). The views expressed are those of the authors and not necessarily those of the NIHR or the Department of Health and Social Care.

## REFERENCES

Aloot, C.B., Vredevoe, D.L., & Brecht, M.L. (1993). Evaluation of high-risk smoking practices used by the homeless. Cancer Nurs, 16, 123–130.

Cane, J., O’Connor, D., & Michie, S. (2012). Validation of the theoretical domains framework for use in behaviour change and implementation research. Implement Sci, 7(37). DOI: 10.1186/1748-5908-7-37

Chen, J.S., Nguyen, A.H., Malesker, M.A., Morrow, L.E. (2016). High-risk smoking behaviors and barriers to smoking cessation among homeless individuals. Respir Care, 1, 640–645. DOI: 10.4187/respcare.04439

Cox, S., Ford, A., Li, J., Best, C., Tyler, A., Robson, D.J., Bauld, L., Hajek, P., Uny, I., Parrott, S.J., & Dawkins, L. (2021). Exploring the uptake and use of electronic cigarettes provided to smokers accessing homeless centres: a four-centre cluster feasibility trial. Public Health Res, 9(7). 10.3310/phr09070

Cox, S., Bauld, L., Brown, R., Carlisle, M., Ford, A., Hajek, P., Li, J., Notley, C., Parrott, S., Pesola, F., Robson, D., Soar, K., Tyler, A., Ward, E., & Dawkins. (2022). Evaluating the effectiveness of e-cigarettes compared with usual care for smoking cessation when offered to smokers at homeless centres: protocol for a multi-centre cluster-randomized controlled trial in Great Britain. Addiction, 117, 2096–2107. 10.1111/add.15851

Dawkins, L., Kimber, C., Puwanesarasa, Y., Soar, K. (2015). First-versus second-generation electronic cigarettes: predictors of choice and effects on urge to smoke and withdrawal symptoms. Addiction, 110, 669–677. 10.1111/add.12807

Dawkins, L., Ford, A., Bauld, L., Balaban, S., Tyler, A., Cox, S. (2019). A cross sectional survey of smoking characteristics and quitting behaviour from a sample of homeless adults in Great Britain. Addict Behav, 95, 35–40. DOI: 10.1016/j.addbeh.2019.02.020

Dawkins, L., Bauld, L., Ford, A., Robson, D., Hajek, P., Parrott, S., Best, C., Li, J., Tyler, A., Uny, I, & Cox, S. (2020). A cluster feasibility trial to explore the uptake and use of e-cigarettes versus usual care offered to smokers attending homeless centres in Great Britain. PLOS ONE, 15(10). 10.1371/journal.pone.0240968

Dawkins, L., Soar, K., Pesola, F., Ford, A., Notley, C., Brown, R., Ward, E., McMillan, L., Robson, D., Varley, A., Mair, C., Lennon, J., Brierley, J., Edwards, A., Hajek, P., Tyler, A., Parrott, S., Li, J., Bauld, L., Gardner, B., Cox, S. (2024). Smoking cessation for people accessing homeless support centres (SCeTCH): comparing the provision of an E-cigarette versus Usual Care in a cluster randomised controlled trial in Great Britain. Pre-print available at SSRN: https://ssrn.com/abstract=4997919 or 10.2139/ssrn.4997919

Flor, L.S., Reitsma, M.B., Gupta, V., Ng, M., Gakidou, E. (2021). The effects of tobacco control policies on global smoking prevalence. Nature Medicine. 27, 239–243. 10.1038/s41591-020-01210-8

Garner, L., & Ratschen, E. (2013). Tobacco smoking, associated risk behaviours, and experience with quitting: A qualitative study with homeless smokers addicted to drugs and alcohol. BMC Public Health, 13, Article 951.

Gilbody, S., Peckham, E., Bailey, D., Arundel, C., Heron, P., Crosland, S., Fairhurst, C., Hewitt, C., Li, J., Parrott, S., Bradshaw, T., Horspool, M., Hughes, E., Hughes, T., Ker, S., Leahy, M., McCloud, T., Osborn, D., Reilly, J., Steare, T., Ballantyne, E., Bidwell, P., Bonner, S., Brennan, D., Callen, T., Carey, A., Colbeck, C., Coton, D., Donaldson, E., Evans, K., Herlihy, H., Khan, W., Nyathi, L., Nyamadzawo, E., Oldknow, H., Phiri, P., Rathod, S., Rea, J., Romain-Hooper, C.B., Smith, K., Stribling, A., & Vickers, C. (2019). Smoking cessation for people with severe mental illness (SCIMITAR+): a pragmatic randomised controlled trial. Lancet Psychiatry. 6(5), 379–390. DOI: 10.1016/S2215-0366(19)30047-1

Guillaumier, A., Skelton, E., Shakeshaft, A., Farrell, M., Tzelepis, F., Walsberger, S., D’Este, C., Paul, C., Dunlop, A., Stirling, R., Fowlie, C., Kelly, P., Oldmeadow, C., Palazzi, K., & Bonevski, B. (2020). Effect of increasing the delivery of smoking cessation care in alcohol and other drug treatment centres: a cluster-randomized controlled trial. Addiction, 115(7), 1345–1355. DOI: 10.1111/add.14911

HM Treasury. (2024). Vaping Products Duty: Consultation Response. https://assets.publishing.service.gov.uk/media/672263b43ce5634f5f6ef582/Vaping_Products_Duty_consultation_response.pdf

Homeless Link. (2022). The Unhealthy State of Homelessness 2022, Homeless Health Needs Audit. https://homelesslink-1b54.kxcdn.com/media/documents/Homeless_Health_Needs_Audit_Report.pdf

Hosseinpoor, A.R., Parker, L.A., Tursan d’Espaignet, E., & Chatterji, S. (2012). Socioeconomic inequality in smoking in low-income and middle-income countries: results from the World Health Survey. PLoS One, 7(8). DOI: 10.1371/journal.pone.0042843

Hummel, K., Brown, J., Willemsen, M.C., West, R., & Kotz, D. (2017). External validation of the Motivation To Stop Scale (MTSS): findings from the International Tobacco Control (ITC) Netherlands Survey. European Journal of Public Health, 27(1), 129–134. 10.1093/eurpub/ckw105

Lindson, N., Butler, A.R., McRobbie, H.J., Bullen, C., Hajek, P., Begh, R., Theodoulou, A., Notley, C., Rigotti, N.A., Turner, T., Livingstone-Banks, J., Morris, T., & Hartmann-Boyce, J. (2024). Electronic cigarettes for smoking cessation. The Cochrane database of systematic reviews, 1. DOI: 10.1002/14651858.CD010216.pub8

Lynch, J.W., Kaplan, G.A., & Salonen, J.T. (1997). Why do poor people behave poorly? Variation in adult health behaviours and psychosocial characteristics by stages of the socioeconomic lifecourse. Soc Sci Med, 44, 809–19. DOI: 10.1016/s0277-9536(96)00191-8

Michie, S., Johnston, M., Abraham, C., Lawton, R., Parker, D., & Walker, A. (2005). Making psychological theory useful for implementing evidence based practice: a consensus approach. Qual Saf Health Care, 14, 26–33. DOI: 10.1136/qshc.2004.011155

Michie, S., Van Stralen, M.M., & West, R. (2011). The behaviour change wheel: a new method for characterising and designing behaviour change interventions. Implement Sci, 6(1). DOI: 10.1186/1748-5908-6-42

Moore, G.F., Audrey, S., Barker, M., Bond, L., Bonell, C., Hardeman, W., Moore, L., O’Cathain, A., Tinati, T., Wight, D., & Baird, J. (2015). Process evaluation of complex interventions: Medical Research Council guidance. BMJ. DOI: 10.1136/bmj.h1258

O’Connell, G., Pritchard, J.D., Prue, C., Thompson, J., Verron, T, Graff, D. & Walele, T. (2019). A randomised, open-label, cross-over clinical study to evaluate the pharmacokinetic profiles of cigarettes and e-cigarettes with nicotine salt formulations in US adult smokers. Intern Emerg Med, 14, 853–61. DOI: 10.1007/s11739-019-02025-3

Office for National Statistics. (2024). Adult smoking habits in the UK: 2023. Population Health Monitoring Group. Retrieved from https://www.ons.gov.uk/peoplepopulationandcommunity/healthandsocialcare/healthandlifeex pectancies/bulletins/adultsmokinghabitsingreatbritain/2023 Accessed December 2, 2024.

Okuyemi, K.S., Caldwell, A.R., Thomas, J.L., Born, W., Richter, K.P., Nollen, N., Braunstein, K., & Ahluwalia, J.S. (2006). Homelessness and smoking cessation: insights from focus groups. Nicotine Tob Res, 8(2), 287–96. DOI: 10.1080/14622200500494971.

Okuyemi, K.S., Goldade, K., Whembolua, G.L., Thomas, J.L., Eischen, S., Sewali, B., Guo, H., Connett, J.E., Grant, J., Ahluwalia, J.S., Resnicow, K., Owen, G., Gelberg, L., & Des Jarlais, D. (2013). Motivational interviewing to enhance nicotine patch treatment for smoking cessation among homeless smokers: a randomized controlled trial. Addiction, 108(6), 1136–44. DOI: 10.1111/add.12140

Notley, C. & Collins, R. (2018). Redefining smoking relapse as recovered social identity – secondary qualitative analysis of relapse narratives. Journal of Substance Use, 23(6), 660–666. 10.1080/14659891.2018.1489009

Notley, C., Ward, E., Dawkins, L., Holland, R., Jakes, S. (2019). Vaping as an alternative to smoking relapse following brief lapse. Drug and Alcohol Review, 38(1), 68–75. 10.1111/dar.12876

Notley, C., Ward, E., Dawkins, L., Holland, R. (2021). User pathways of e-cigarette use to support long term tobacco smoking relapse prevention: a qualitative analysis. Addiction, 116(3), 596–605. DOI: 10.1111/add.15226.

Papadakis, S. & McEwen, A. (2021). Very brief advice on smoking PLUS (VBA+). National Centre for Smoking Cessation and Training (NCSCT). Retrieved from https://www.ncsct.co.uk/publication_VBA+.php Accessed December 2 2024

Porter, M., Harvey, J., Gavin, J.K., Carpenter, M.J., Cummings, M., Pope, C., & Diaz, V.A. (2017). Qualitative Study to Assess Factors Supporting Tobacco Use in A Homeless Population. AIMS Medical Science, 4(1), 83–98. DOI: 10.3934/medsci.2017.1.83

Pratt, R., Pernat, C., Kerandi, L., Kmiecik, A., Strobel-Ayres, C., Joseph, A., Everson Rose, S.A., Luo, X., Cooney, N., Thomas, J., & Okuyemi, K. (2019). “It’s a hard thing to manage when you’re homeless”: the impact of the social environment on smoking cessation for smokers experiencing homelessness. BMC Public Health, 19. 10.1186/s12889-019-6987-7

Sallis, J., Owen, N., & Fisher, E. (2008). Ecological models of health behaviour. In: Glanz, K., Rimer, B., & Viswanath, K., (Eds.), Health behavior and health education: theory, research, and practice. 4th ed. (pp. 465–482). Jossey-Bass.

Soar, K., Dawkins, L., Robson, D., & Cox, S. (2020). Smoking amongst adults experiencing homelessness: a systematic review of prevalence rates, interventions and the barriers and facilitators to quitting and staying quit. Journal of Smoking Cessation, 15(2), 94–108. DOI:10.1017/jsc.2020.11

Stewart, H. C., Stevenson, T. N., Bruce, J. S., Greenberg, B., & Chamberlain, L. J. (2015). Attitudes Toward Smoking Cessation Among Sheltered Homeless Parents. Journal of community health, 40(6), 1140–1148. 10.1007/s10900-015-0040-2

Taylor, M., Mackay, K., Murphy, J., McIntosh, A., McIntosh, C., Anderson, S., & Welch, K. (2012). Quantifying the RR of harm to self and others from substance misuse: results from a survey of clinical experts across Scotland. BMJ open, 2(4). 10.1136/bmjopen-2011-000774

Vijayaraghavan, M., Elser, H., Frazer, K., Lindson, N., & Apollonio, D. (2020). Interventions to reduce tobacco use in people experiencing homelessness. The Cochrane database of systematic reviews, 12(12). 10.1002/14651858.CD013413.pub2

Voos, N., Kaiser, L., Mahoney, M. C., Bradizza, C. M., Kozlowski, L. T., Benowitz, N. L., O’Connor, R. J., & Goniewicz, M. L. (2019). Randomized within-subject trial to evaluate smokers’ initial perceptions, subjective effects and nicotine delivery across six vaporized nicotine products. Addiction, 114(7), 1236–1248. 10.1111/add.14602

Yong, H. H., Borland, R., Cummings, K. M., Gravely, S., Thrasher, J. F., McNeill, A., Hitchman, S., Greenhalgh, E., Thompson, M. E., & Fong, G. T. (2019). Reasons for regular vaping and for its discontinuation among smokers and recent ex-smokers: findings from the 2016 ITC Four Country Smoking and Vaping Survey. Addiction, 114, 35–48. 10.1111/add.14593

