## Supplementary figures and images for "Exploring how an e-cigarette intervention influenced tobacco smoking behaviour in people accessing homelessness services: findings from the SCeTCH trial process evaluation"

### Supplementary figure 1 SCeTCH trial logic model

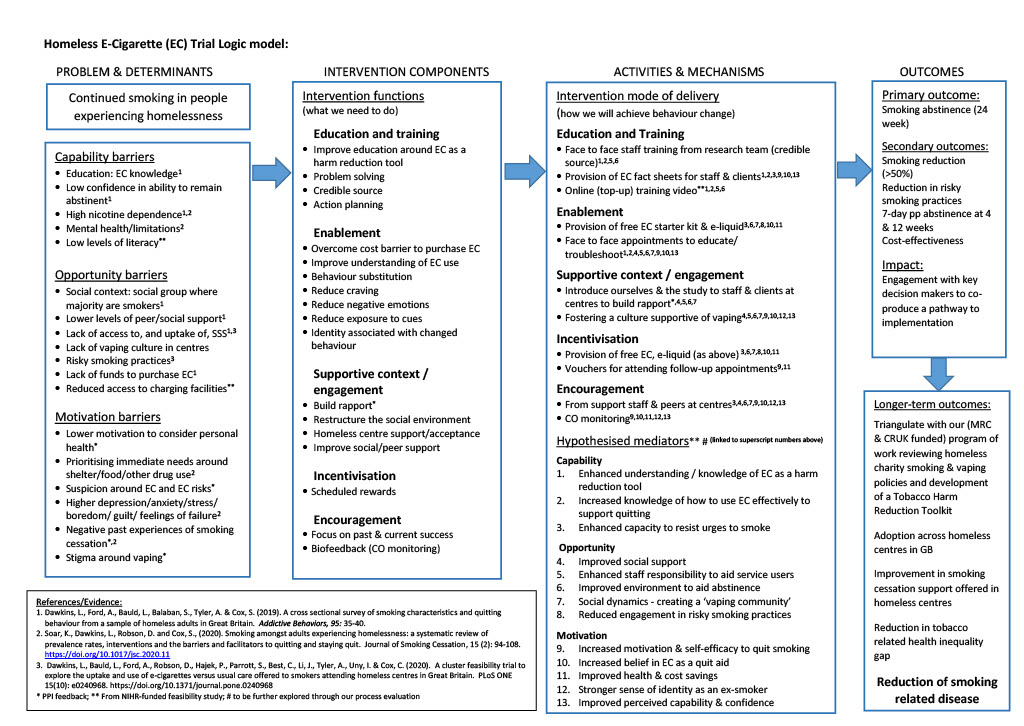
